# Type 1 and type 2 diabetes mellitus: Clinical outcomes due to COVID-19. Protocol of a systematic literature review

**DOI:** 10.1101/2022.07.11.22277475

**Authors:** Juan Pablo Pérez Bedoya, Alejandro Mejía Muñoz, Noël Christopher Barengo, Paula Andrea Diaz Valencia

## Abstract

**Introduction:** Diabetes has been associated with an increased risk of complications in patients with COVID-19. Most studies do not differentiate between patients with type 1 and type 2 diabetes, which correspond to two pathophysiological distinct diseases that could represent different degrees of clinical compromise.

**Objective:** To identify if there are differences in the clinical outcomes of patients with COVID-19 and diabetes (type 1 and type 2) compared to patients with COVID-19 without diabetes.

**Methods:** Observational studies of patients with COVID-19 and diabetes (both type 1 and type 2) will be included without restriction of geographic region, gender or age, whose outcome is hospitalization, admission to intensive care unit or mortality compared to patients without diabetes. Two authors will independently perform selection, data extraction, and quality assessment, and a third reviewer will resolve discrepancies. The data will be synthesized regarding the sociodemographic and clinical characteristics of patients with diabetes and without diabetes accompanied by the measure of association for the outcomes. The data will be synthesized regarding the sociodemographic and clinical characteristics of patients with diabetes and without diabetes accompanied by the measure of association for the outcomes.

**Expected results:** Update the evidence regarding the risk of complications in diabetic patients with COVID-19 and in turn synthesize the information available regarding type 1 and type 2 diabetes mellitus, to provide keys to a better understanding of the pathophysiology of diabetics.

**Systematic review registry:** This study was registered at the International Prospective Registry for Systematic Reviews (PROSPERO) - CRD42021231942.

## Introduction

The Severe Acute Respiratory Syndrome Coronavirus type 2 (SARS-CoV-2), the causal viral agent of coronavirus disease 2019 (COVID-19), currently has the world in one of the greatest public health crises of recent times since its appearance at the end of 2019 in the city of Wuhan, China [1]. The infection has a mild or even asymptomatic course in most cases, but in elderly patients (over 60 years-of-age) and in those with pre-existing chronic comorbidities, it can progress severe complications such as pneumonia, acute respiratory distress (ARDS) with hyperinflammatory involvement and multi-organ failure, leading in some cases to death [2].

Different studies have reported that patients diagnosed with diabetes who suffer from COVID-19 disease have higher morbidity and mortality compared with people without diabetes [3]. An analysis by Gude Sampedro et al using prognostic models found that diabetic patients had greater odds of being hospitalized (OR 1.43; 95% CI: 1.18 to 1.73), admitted to the intensive care unit (OR 1.61; 95% CI: 1.12 to 2.31) and dying from COVID-19 (OR 1.79; 95% CI %: 1.38 to 2.32) compared with patients without diabetes [4]. However, it is difficult to establish whether diabetes alone directly contributed to the increase likelihood of complications.

Several studies using secondary data have emerged during the course of the pandemic that seek to determine the association of diabetes with mortality and other clinical outcomes in patients with COVID-19, such as, for example, a meta-analysis carried out by Shang et al. of severe infection and mortality from COVID-19 in diabetic patients compared with those without diabetes. They reported that patients with COVID-19 and diabetes had higher odds of serious infection (OR = 2.38, 95% CI: 2.05 to 2.78) and mortality (OR = 2, 21, 95% CI: 1.83 to 2.66) than patients without diabetes [5]. Despite the fact that there are several primary studies that attempt to explain the association between diabetes and COVID-19, most studies lack epidemiological rigor in the design and methodology used [6]. In addition, many of them did not distinguish between type 1 and type 2 diabetes, which are two very different conditions with different clinical development and pathophysiological mechanisms [7]. This may lead to different degrees of clinical complications from COVID-19. Currently, there is a gap in knowledge about the complications in patients with COVID-19 according to the type of diabetes. Moreover, only limited information exist how COVID-19 affects type 1 patients [8,9].

The objective of this systematic literature review will be to identify whether there are differences in the clinical outcomes of both type 1 and type 2 diabetes patients diagnosed with COVID-19 compared with patients with COVID-19 without a diagnosis of diabetes. This study will provide scientific evidence regarding the risk of complications in diabetic patients with COVID-19 and, in turn, synthesize the available information regarding to type 1 and type 2 diabetes.

## Methods/Design

### Study design

This systematic literature review protocol was prepared according to the Preferred Reporting Elements for Systematic Review and Meta-Analysis Protocols (PRISMA-P) [10] (S1 Appendix). The results of the final systematic review will be reported according to the preferred reporting items for systematic reviews and meta-analyses (PRISMA 2020) [11,12]. In the event of significant deviations from this protocol, they will be reported and published with the results of the review.

### Eligibility Criteria

#### Participants (population)

Patients with a confirmed diagnosis of COVID-19 without restriction of geographic region, gender or age of the participants, in which the diagnostic algorithm used to confirm SARS-CoV-2 infection is adequately presented, as it is the case of standardized molecular biology laboratory methods such as RT-PCR (real-time reverse transcriptase polymerase chain reaction) and/or antigen test for the detection of virus surface proteins, or failing that, diagnosis clinical or epidemiological based on international guidelines and recommendations by the WHO or/and the Center for Disease Control and Prevention (CDC).

#### Exposure

Patients with COVID-19 and concomitant diagnosis of unspecified diabetes mellitus, differentiated into type 1 diabetes mellitus or type 2 diabetes mellitus, without restriction of geographic region, gender, or age of the patients, who present definition of clinical criteria and /or paraclinical tests used by researchers to classify patients according to their diabetes status.

#### Comparator

Patients with COVID-19 who do not have a concomitant diagnosis of diabetes mellitus.

#### Outcome

The main endpoint is all-cause mortality (according to the definitions of each primary study) and the secondary outcomes are hospitalization and admission to the ICU, where the authors specify a clear definition based on clinical practice guidelines and provide a well-defined criteria for patient outcomes.

#### Type of study

Primary observational original research studies (prospective or retrospective cohort, case-control design, and cross-sectional studies) will be included in this systematic review.

### Exclusion criteria

Clinical trials, editorials, letters to the editor, reviews, case reports, case series, narrative reviews or systematic reviews and meta-analyses, as well as research in the field of basic sciences based on experimental laboratory models, will be excluded. Similarly, original research articles that only include other types of diabetes such as monogenic diabetes, gestational diabetes, latent autoimmune diabetes in adults, ketosis-prone diabetes, among others, or articles with a publication status prior to review will be excluded.

### Information sources and search strategy

For the preparation of the search strategy, the recommendations of the PRISMA-S guide [13] will be adopted. Relevant articles will be identified by electronic search applying the equation previously developed by the researchers and validated by an expert librarian (S2 Appendix). The following electronic bibliographic databases will be used: MEDLINE, EMBASE, LILACS, OVID MEDLINE and WHO (COVID-19 Global literature on coronavirus disease) with a publication date from December 2019 to March 20, 2022, without language restriction. To identify other potentially eligible studies, the references of relevant publications will be reviewed to perform a snowball manual search.

### Study selection process

Two investigators will independently assess all the titles and abstracts of the retrieved articles using the free access Rayyan software® [14]. Disagreement will be resolved in the first instance through discussion and in a second instance through a third reviewer. Articles that meet the eligibility criteria will be included. The full text will be read independently by two researchers. Discrepancies will be resolved through discussion or a third reviewer. The process of identification, screening and inclusion of primary studies will be described and presented using the flow chart recommended by the PRISMA statement in its latest version 2020 [11,12].

### Data collection and extraction

Standardized and validated forms will be used to collect the data extracted from the primary studies, accompanied by a detailed instruction manual to specify the guiding questions, and avoid the introduction of bias. This process will be carried out by two researchers independently. A third investigator will verify the extracted data to ensure the accuracy of the records. The authors of the primary studies will be contacted to resolve any questions that may arise. The reviewers will resolve disagreements through discussion and one of the two referees will adjudicate the discrepancies presented through discussion and consensus.

In specific terms, the following data will be collected both for the primary studies that report diabetes and COVID-19 and for those that differentiate between DMT1 and DMT2: author, year and country where the study was carried out; study design; general characteristics of the population, sample size, demographic data of the participants (sex, age, ethnicity), percentage of patients with diabetes, percentage of patients with type 1 and/or type 2 diabetes, percentage of patients without diabetes, frequency of comorbidities in diabetics and non-diabetics, percentage of diabetic and non-diabetic patients who presented the outcomes (hospitalization, ICU admission and mortality) and association measures reported for the outcomes. Data extraction will be done using a Microsoft Excel 365 ® spreadsheets.

### Quality evaluation

The study quality assessment tool provided by the National Institutes of Health (NIH) [15] will be used for observational studies such as cohort, case-control, and cross-sectional. Two tools will be sued: one for cohort and cross-sectional studies (14 questions/domains) and one for case-control studies (12 questions/domains). These tools are aimed at detecting elements that allow evaluation of possible methodological problems, including sources of bias (for example, patient selection, performance, attrition and detection), confounding, study power, the strength of causality in the association between interventions and outcomes, among other factors. The different tools that will be used reflect a score of “1” or “0” depending on the answer “yes” or “no”, respectively for each question or domain evaluated, or failing that, the indeterminate criterion option. For observational cohort studies, which consist of 14 risk of bias assessment domains, the studies will be classified as having good quality if they obtain ≥10 points, of fair quality if they obtain 8 to 9 points, and of poor quality if they obtain less than 8 points. On the other hand, in the case of case-control studies that consist of 12 bias risk assessment domains, the studies will be classified as good quality if they obtained ≥8 points, regular quality if they obtained 6 to 7 points and of poor quality if they obtained less than 6 points. However, the internal discussion between the research team will always be considered as the primary quality criterion.

### Data synthesis

A narrative synthesis with summary tables will be carried out according to the recommendations adapted from the Synthesis Without Meta-analysis (SWiM) guide to describe in a structured way the methods used, and the findings found in the primary studies, as well as the criteria for grouping of the studies [16]. A narrative synthesis will be presented in two sections, one for patients with COVID-19 and diabetes and another for patients with COVID-19 and type 1 or type 2 diabetes.

Assessment of clinical and methodological heterogeneity will determine the feasibility of the meta-analysis. Possible sources of heterogeneity identified are the clinical characteristics of the study population, the criteria used to define the outcomes in the groups of patients, the time period of the pandemic in which the study was carried out, and the availability of measurement and control for potential confounding factors. For this reason, it is established a priori that this diversity of findings will make it difficult to carry out an adequate meta-analysis [17]. However, if meta-analysis is considered feasible, the random effects model will be used due to the high probability of heterogeneity between studies. Statistical heterogeneity will be assessed using the X^2^ test and the I^2^ statistic, and publication bias assessed using funnel plots if there are sufficient (>10) studies [18].

### Exploratory ecological analysis

An exploratory ecological analysis of the association between the frequency of clinical outcomes of diabetic patients with COVID-19 and the indicators related to the health care dimension, reported for the different countries analyzed by means of the correlation coefficient, will be carried out. The open public databases of the World Bank (WB) [19], the World Health Organization (WHO) [20] and Our World In Data [21] will be used to extract population indicators related to health care, among those prioritized, universal health coverage, hospital beds per 1,000 people, doctors per 1,000 people, current health spending as a percentage of gross domestic product (GDP), percentage of complete vaccination coverage for COVID-19.

## Discussion

Since the first epidemiological and clinical reports were released from the city of Wuhan regarding the clinical characteristics of patients with COVID-19, a high incidence of chronic non-communicable diseases has been observed in Covid-19 patients. Current scientific evidence has shown that certain comorbidities increase the risk for hospitalization, severity of illness or death from COVID-19, such as hypertension, cardiovascular disease, chronic kidney disease, chronic respiratory disease, diabetes, among others [22].

One of the main chronic comorbidities affected by the COVID-19 pandemic is diabetes. Multivariate analysis of several observational epidemiological studies have revealed that COVID-19 patients with diabetes were at increased risk of hospitalization, ICU admission, and mortality compared with patients without diabetes [4].

For this reason, it is expected that this systematic literature review will provide scientific support regarding the outcomes and complications that patients diagnosed with COVID-19 with type 1 or type 2 diabetes present compared with patients without diabetes. This information will be useful for healthcare personnel, public health professionals and epidemiologists involved in patient care or decision making, generating epidemiological evidence. Thus, highlighting the decisive role of epidemiological research in the context of the pandemic, especially in the field of diabetes epidemiology may improve comprehensive management and care of diabetic patients. This study may also provide important information that can be used to update of clinical practice guidelines.

## The status of the study

The study is in the selection phase of the records by applying the eligibility criteria to the titles and abstracts. Completion of the project is expected in September 2022 with the publication of the results.

## Conclusions

This report describes the systematic review protocol that will be utilized to update the evidence regarding the risk of complications in diabetic patients with COVID-19 and in turn synthesize the information available regarding DM1 and DM2, to provide keys to a better understanding of the pathophysiology of diabetics.

## Data Availability

All relevant data from this study will be made available upon study completion.

## Acknowledgments

This research was developed within the “Repository for the surveillance of risk factors for chronic diseases in Colombia, the Caribbean and the Americas” framework and financially supported by a grant of Colciencias 844 (grant number 111584467754).

## Supporting information

**S1 Appendix**. PRISMA-P (Preferred Reporting Items for Systematic review and Meta-Analysis Protocols) 2015 checklist: Recommended items to address in a systematic review protocol.

**S2 Appendix**. Search string details for each database.

