## Supplementary material for "Type 1 and type 2 diabetes mellitus: Clinical outcomes due to COVID-19. Protocol of a systematic literature review": S2 Appendix

**Appendix 1. Search equations for the different sources of information or electronic bibliographic databases**

The following search equations were constructed by the research team and subsequently validated by a librarian who is an expert in searching for information related to the health area. The following results correspond to the closing of the search carried out on March 20, 2022.

**PUBMED:**

**#1:** (((((((((((((((((((((((((((((((((((((((((((((((((((((((((((((((((((((((((((((((((((((((((((COVID-19[MeSH Terms]) OR (COVID-19[Title/Abstract])) OR (SARS-CoV-2[MeSH Terms])) OR (SARS-CoV-2[Title/Abstract])) OR (SARS-CoV-2 Infection[MeSH Terms])) OR (SARS-CoV-2 Infection[Title/Abstract])) OR (Infection, SARS-CoV-2[MeSH Terms])) OR (Infection, SARS-CoV-2[Title/Abstract])) OR (SARS-CoV-2 Infections[MeSH Terms])) OR (SARS-CoV-2 Infections[Title/Abstract])) OR (2019 Novel Coronavirus Disease[MeSH Terms])) OR (2019 Novel Coronavirus Disease[Title/Abstract])) OR (2019 Novel Coronavirus Infection[MeSH Terms])) OR (2019 Novel Coronavirus Infection[Title/Abstract])) OR (2019-nCoV Disease[MeSH Terms])) OR (2019-nCoV Disease[Title/Abstract])) OR (2019-nCoV Diseases[MeSH Terms])) OR (2019-nCoV Diseases[Title/Abstract])) OR (Disease, 2019-nCoV[MeSH Terms])) OR (Disease, 2019-nCoV[Title/Abstract])) OR (COVID-19 Virus Infection[MeSH Terms])) OR (COVID-19 Virus Infection[Title/Abstract])) OR (COVID-19 Virus Infections[MeSH Terms])) OR (COVID-19 Virus Infections[Title/Abstract])) OR (Infection, COVID-19 Virus[MeSH Terms])) OR (Infection, COVID-19 Virus[Title/Abstract])) OR (Virus Infection, COVID-19[MeSH Terms])) OR (Virus Infection, COVID-19[Title/Abstract])) OR (Coronavirus Disease 2019[MeSH Terms])) OR (Coronavirus Disease 2019[Title/Abstract])) OR (Disease 2019, Coronavirus[MeSH Terms])) OR (Disease 2019, Coronavirus[Title/Abstract])) OR (Coronavirus Disease-19[MeSH Terms])) OR (Coronavirus Disease-19[Title/Abstract])) OR (Severe Acute Respiratory Syndrome Coronavirus 2 Infection[MeSH Terms])) OR (Severe Acute Respiratory Syndrome Coronavirus 2 Infection[Title/Abstract])) OR (SARS Coronavirus 2 Infection[MeSH Terms])) OR (SARS Coronavirus 2 Infection[Title/Abstract])) OR (COVID-19 Virus Disease[MeSH Terms])) OR (COVID-19 Virus Disease[Title/Abstract])) OR (COVID-19 Virus Diseases[MeSH Terms])) OR (COVID-19 Virus Diseases[Title/Abstract])) OR (Disease, COVID-19 Virus[MeSH Terms])) OR (Disease, COVID-19 Virus[Title/Abstract])) OR (Virus Disease, COVID-19[MeSH Terms])) OR (Virus Disease, COVID-19[Title/Abstract])) OR (2019-nCoV Infection[MeSH Terms])) OR (2019-nCoV Infection[Title/Abstract])) OR (2019-nCoV Infections[MeSH Terms])) OR (2019-nCoV Infections[Title/Abstract])) OR (Infection, 2019-nCoV[MeSH Terms])) OR (Infection, 2019-nCoV[Title/Abstract])) OR (COVID-19 Pandemic[MeSH Terms])) OR (COVID-19 Pandemic[Title/Abstract])) OR (Pandemic, COVID-19[MeSH Terms])) OR (Pandemic, COVID-19[Title/Abstract])) OR (COVID-19 Pandemics[MeSH Terms])) OR (COVID-19 Pandemics[Title/Abstract])) OR (SARS Coronavirus 2[MeSH Terms])) OR (SARS Coronavirus 2[Title/Abstract])) OR (Coronavirus 2, SARS[MeSH Terms])) OR (Coronavirus 2, SARS[Title/Abstract])) OR (Coronavirus Disease 2019 Virus[MeSH Terms])) OR (Coronavirus Disease 2019 Virus[Title/Abstract])) OR (2019 Novel Coronavirus[MeSH Terms])) OR (2019 Novel Coronavirus[Title/Abstract])) OR (2019 Novel Coronaviruses[MeSH Terms])) OR (2019 Novel Coronaviruses[Title/Abstract])) OR (Coronavirus, 2019 Novel[MeSH Terms])) OR (Coronavirus, 2019 Novel[Title/Abstract])) OR (Novel Coronavirus, 2019[MeSH Terms])) OR (Novel Coronavirus, 2019[Title/Abstract])) OR (SARS-CoV-2 Virus[MeSH Terms])) OR (SARS-CoV-2 Virus[Title/Abstract])) OR (SARS-CoV-2 Viruses[MeSH Terms])) OR (SARS-CoV-2 Viruses[Title/Abstract])) OR (Virus, SARS-CoV-2[MeSH Terms])) OR (Virus, SARS-CoV-2[Title/Abstract])) OR (2019-nCoV[MeSH Terms])) OR (2019-nCoV[Title/Abstract])) OR (COVID-19 Virus[MeSH Terms])) OR (COVID-19 Virus[Title/Abstract])) OR (COVID-19 Viruses[MeSH Terms])) OR (COVID-19 Viruses[Title/Abstract])) OR (Virus, COVID-19[MeSH Terms])) OR (Virus, COVID-19[Title/Abstract])) OR (Wuhan Coronavirus[MeSH Terms])) OR (Wuhan Coronavirus[Title/Abstract])) OR (Coronavirus, Wuhan[MeSH Terms])) OR (Coronavirus, Wuhan[Title/Abstract])) OR (Severe Acute Respiratory Syndrome Coronavirus 2[MeSH Terms])) OR (Severe Acute Respiratory Syndrome Coronavirus 2[Title/Abstract])

**#2:** (((((((((((((((((((((((((((((((((((((((Diabetes Mellitus[MeSH Terms]) OR (Diabetes Mellitus[Title/Abstract])) OR (Diabetes[Title/Abstract])) OR (Diabetic[Title/Abstract])) OR (Diabetes Mellitus, Type 1[MeSH Terms])) OR (Diabetes Mellitus, Type 1[Title/Abstract])) OR (Diabetes Mellitus, Insulin-Dependent[Title/Abstract])) OR (Diabetes Mellitus, Insulin Dependent[Title/Abstract])) OR (Insulin-Dependent Diabetes Mellitus[Title/Abstract])) OR (Diabetes Mellitus, Juvenile-Onset[Title/Abstract])) OR (Diabetes Mellitus, Juvenile Onset[Title/Abstract])) OR (Juvenile-Onset Diabetes Mellitus[Title/Abstract])) OR (Type 1 Diabetes Mellitus[Title/Abstract])) OR (Diabetes Mellitus, Insulin-Dependent, 1[Title/Abstract])) OR (Insulin-Dependent Diabetes Mellitus 1[Title/Abstract])) OR (Type 1 Diabetes[Title/Abstract])) OR (Diabetes, Type 1[Title/Abstract])) OR (Diabetes Mellitus, Type I[Title/Abstract])) OR (Diabetes, Autoimmune[Title/Abstract])) OR (Autoimmune Diabetes[Title/Abstract])) OR (IDDM[Title/Abstract])) OR (T1DM[Title/Abstract])) OR (Diabetes Mellitus, Type 2[MeSH Terms])) OR (Diabetes Mellitus, Type 2[Title/Abstract])) OR (Diabetes Mellitus, Noninsulin-Dependent[Title/Abstract])) OR (Diabetes Mellitus, Non Insulin Dependent[Title/Abstract])) OR (Diabetes Mellitus, Non-Insulin-Dependent[Title/Abstract])) OR (Non-Insulin-Dependent Diabetes Mellitus[Title/Abstract])) OR (Diabetes Mellitus, Type II[Title/Abstract])) OR (NIDDM[Title/Abstract])) OR (Diabetes Mellitus, Noninsulin Dependent[Title/Abstract])) OR (Type 2 Diabetes Mellitus[Title/Abstract])) OR (Noninsulin-Dependent Diabetes Mellitus[Title/Abstract])) OR (Noninsulin Dependent Diabetes Mellitus[Title/Abstract])) OR (Type 2 Diabetes[Title/Abstract])) OR (Diabetes, Type 2[Title/Abstract])) OR (Diabetes Mellitus, Adult-Onset[Title/Abstract])) OR (Adult-Onset Diabetes Mellitus[Title/Abstract])) OR (Diabetes Mellitus, Adult Onset[Title/Abstract])) OR (T2DM[Title/Abstract])

**#3:** ((((((((((((((((((((((((((((((((((((((((((((((((((((((((((((((((((((((((((((((((((((((((((((COVID-19[MeSH Terms]) OR (COVID-19[Title/Abstract])) OR (SARS-CoV-2[MeSH Terms])) OR (SARS-CoV-2[Title/Abstract])) OR (SARS-CoV-2 Infection[MeSH Terms])) OR (SARS-CoV-2 Infection[Title/Abstract])) OR (Infection, SARS-CoV-2[MeSH Terms])) OR (Infection, SARS-CoV-2[Title/Abstract])) OR (SARS-CoV-2 Infections[MeSH Terms])) OR (SARS-CoV-2 Infections[Title/Abstract])) OR (2019 Novel Coronavirus Disease[MeSH Terms])) OR (2019 Novel Coronavirus Disease[Title/Abstract])) OR (2019 Novel Coronavirus Infection[MeSH Terms])) OR (2019 Novel Coronavirus Infection[Title/Abstract])) OR (2019-nCoV Disease[MeSH Terms])) OR (2019-nCoV Disease[Title/Abstract])) OR (2019-nCoV Diseases[MeSH Terms])) OR (2019-nCoV Diseases[Title/Abstract])) OR (Disease, 2019-nCoV[MeSH Terms])) OR (Disease, 2019-nCoV[Title/Abstract])) OR (COVID-19 Virus Infection[MeSH Terms])) OR (COVID-19 Virus Infection[Title/Abstract])) OR (COVID-19 Virus Infections[MeSH Terms])) OR (COVID-19 Virus Infections[Title/Abstract])) OR (Infection, COVID-19 Virus[MeSH Terms])) OR (Infection, COVID-19 Virus[Title/Abstract])) OR (Virus Infection, COVID-19[MeSH Terms])) OR (Virus Infection, COVID-19[Title/Abstract])) OR (Coronavirus Disease 2019[MeSH Terms])) OR (Coronavirus Disease 2019[Title/Abstract])) OR (Disease 2019, Coronavirus[MeSH Terms])) OR (Disease 2019, Coronavirus[Title/Abstract])) OR (Coronavirus Disease-19[MeSH Terms])) OR (Coronavirus Disease-19[Title/Abstract])) OR (Severe Acute Respiratory Syndrome Coronavirus 2 Infection[MeSH Terms])) OR (Severe Acute Respiratory Syndrome Coronavirus 2 Infection[Title/Abstract])) OR (SARS Coronavirus 2 Infection[MeSH Terms])) OR (SARS Coronavirus 2 Infection[Title/Abstract])) OR (COVID-19 Virus Disease[MeSH Terms])) OR (COVID-19 Virus Disease[Title/Abstract])) OR (COVID-19 Virus Diseases[MeSH Terms])) OR (COVID-19 Virus Diseases[Title/Abstract])) OR (Disease, COVID-19 Virus[MeSH Terms])) OR (Disease, COVID-19 Virus[Title/Abstract])) OR (Virus Disease, COVID-19[MeSH Terms])) OR (Virus Disease, COVID-19[Title/Abstract])) OR (2019-nCoV Infection[MeSH Terms])) OR (2019-nCoV Infection[Title/Abstract])) OR (2019-nCoV Infections[MeSH Terms])) OR (2019-nCoV Infections[Title/Abstract])) OR (Infection, 2019-nCoV[MeSH Terms])) OR (Infection, 2019-nCoV[Title/Abstract])) OR (COVID-19 Pandemic[MeSH Terms])) OR (COVID-19 Pandemic[Title/Abstract])) OR (Pandemic, COVID-19[MeSH Terms])) OR (Pandemic, COVID-19[Title/Abstract])) OR (COVID-19 Pandemics[MeSH Terms])) OR (COVID-19 Pandemics[Title/Abstract])) OR (SARS Coronavirus 2[MeSH Terms])) OR (SARS Coronavirus 2[Title/Abstract])) OR (Coronavirus 2, SARS[MeSH Terms])) OR (Coronavirus 2, SARS[Title/Abstract])) OR (Coronavirus Disease 2019 Virus[MeSH Terms])) OR (Coronavirus Disease 2019 Virus[Title/Abstract])) OR (2019 Novel Coronavirus[MeSH Terms])) OR (2019 Novel Coronavirus[Title/Abstract])) OR (2019 Novel Coronaviruses[MeSH Terms])) OR (2019 Novel Coronaviruses[Title/Abstract])) OR (Coronavirus, 2019 Novel[MeSH Terms])) OR (Coronavirus, 2019 Novel[Title/Abstract])) OR (Novel Coronavirus, 2019[MeSH Terms])) OR (Novel Coronavirus, 2019[Title/Abstract])) OR (SARS-CoV-2 Virus[MeSH Terms])) OR (SARS-CoV-2 Virus[Title/Abstract])) OR (SARS-CoV-2 Viruses[MeSH Terms])) OR (SARS-CoV-2 Viruses[Title/Abstract])) OR (Virus, SARS-CoV-2[MeSH Terms])) OR (Virus, SARS-CoV-2[Title/Abstract])) OR (2019-nCoV[MeSH Terms])) OR (2019-nCoV[Title/Abstract])) OR (COVID-19 Virus[MeSH Terms])) OR (COVID-19 Virus[Title/Abstract])) OR (COVID-19 Viruses[MeSH Terms])) OR (COVID-19 Viruses[Title/Abstract])) OR (Virus, COVID-19[MeSH Terms])) OR (Virus, COVID-19[Title/Abstract])) OR (Wuhan Coronavirus[MeSH Terms])) OR (Wuhan Coronavirus[Title/Abstract])) OR (Coronavirus, Wuhan[MeSH Terms])) OR (Coronavirus, Wuhan[Title/Abstract])) OR (Severe Acute Respiratory Syndrome Coronavirus 2[MeSH Terms])) OR (Severe Acute Respiratory Syndrome Coronavirus 2[Title/Abstract])) AND ((((((((((((((((((((((((((((((((((((((((Diabetes Mellitus[MeSH Terms]) OR (Diabetes Mellitus[Title/Abstract])) OR (Diabetes[Title/Abstract])) OR (Diabetic[Title/Abstract])) OR (Diabetes Mellitus, Type 1[MeSH Terms])) OR (Diabetes Mellitus, Type 1[Title/Abstract])) OR (Diabetes Mellitus, Insulin-Dependent[Title/Abstract])) OR (Diabetes Mellitus, Insulin Dependent[Title/Abstract])) OR (Insulin-Dependent Diabetes Mellitus[Title/Abstract])) OR (Diabetes Mellitus, Juvenile-Onset[Title/Abstract])) OR (Diabetes Mellitus, Juvenile Onset[Title/Abstract])) OR (Juvenile-Onset Diabetes Mellitus[Title/Abstract])) OR (Type 1 Diabetes Mellitus[Title/Abstract])) OR (Diabetes Mellitus, Insulin-Dependent, 1[Title/Abstract])) OR (Insulin-Dependent Diabetes Mellitus 1[Title/Abstract])) OR (Type 1 Diabetes[Title/Abstract])) OR (Diabetes, Type 1[Title/Abstract])) OR (Diabetes Mellitus, Type I[Title/Abstract])) OR (Diabetes, Autoimmune[Title/Abstract])) OR (Autoimmune Diabetes[Title/Abstract])) OR (IDDM[Title/Abstract])) OR (T1DM[Title/Abstract])) OR (Diabetes Mellitus, Type 2[MeSH Terms])) OR (Diabetes Mellitus, Type 2[Title/Abstract])) OR (Diabetes Mellitus, Noninsulin-Dependent[Title/Abstract])) OR (Diabetes Mellitus, Non Insulin Dependent[Title/Abstract])) OR (Diabetes Mellitus, Non-Insulin-Dependent[Title/Abstract])) OR (Non-Insulin-Dependent Diabetes Mellitus[Title/Abstract])) OR (Diabetes Mellitus, Type II[Title/Abstract])) OR (NIDDM[Title/Abstract])) OR (Diabetes Mellitus, Noninsulin Dependent[Title/Abstract])) OR (Type 2 Diabetes Mellitus[Title/Abstract])) OR (Noninsulin-Dependent Diabetes Mellitus[Title/Abstract])) OR (Noninsulin Dependent Diabetes Mellitus[Title/Abstract])) OR (Type 2 Diabetes[Title/Abstract])) OR (Diabetes, Type 2[Title/Abstract])) OR (Diabetes Mellitus, Adult-Onset[Title/Abstract])) OR (Adult-Onset Diabetes Mellitus[Title/Abstract])) OR (Diabetes Mellitus, Adult Onset[Title/Abstract])) OR (T2DM[Title/Abstract]))

**#4:** ((((((((((((((((((((((((((((((((((((((Hospital*[MeSH Terms]) OR (Hospital*[Title/Abstract])) OR (Patient Admission[MeSH Terms])) OR (Patient Admission[Title/Abstract])) OR (Admission, Patient[Title/Abstract])) OR (Admissions, Patient[Title/Abstract])) OR (Patient Admissions[Title/Abstract])) OR (Stay, Hospital[Title/Abstract])) OR (Stays, Hospital[Title/Abstract])) OR (Intensive Care Units[MeSH Terms])) OR (Intensive Care Units[Title/Abstract])) OR (Intensive Care Unit[Title/Abstract])) OR (Unit, Intensive Care[Title/Abstract])) OR (ICU Intensive Care Units[Title/Abstract])) OR (ICU admission[Title/Abstract])) OR (ICU[Title/Abstract])) OR (Critical Care[MeSH Terms])) OR (Critical Care[Title/Abstract])) OR (Care, Critical[Title/Abstract])) OR (Intensive Care[Title/Abstract])) OR (Care, Intensive[Title/Abstract])) OR (Critical Care Unit[Title/Abstract])) OR (Critical Care Units[Title/Abstract])) OR (Intensive Therapy Unit[Title/Abstract])) OR (Intensive Treatment Unit[Title/Abstract])) OR (Critical Care Outcomes[MeSH Terms])) OR (Critical Care Outcomes[Title/Abstract])) OR (Care Outcome, Critical[Title/Abstract])) OR (Care Outcomes, Critical[Title/Abstract])) OR (Critical Care Outcome[Title/Abstract])) OR (Outcome, Critical Care[Title/Abstract])) OR (Outcomes, Critical Care[Title/Abstract])) OR (Mortal*[MeSH Terms])) OR (Mortal*[Title/Abstract])) OR (Death[Title/Abstract])) OR (Death Rate[Title/Abstract])) OR (Death Rates[Title/Abstract])) OR (Mortality Rate[Title/Abstract])) OR (Rate, Mortality[Title/Abstract])

**#5:** (((((((((((((((((((((((((((((((((((((((((((((((((((((((((((((((((((((((((((((((((((((((((((((COVID-19[MeSH Terms]) OR (COVID-19[Title/Abstract])) OR (SARS-CoV-2[MeSH Terms])) OR (SARS-CoV-2[Title/Abstract])) OR (SARS-CoV-2 Infection[MeSH Terms])) OR (SARS-CoV-2 Infection[Title/Abstract])) OR (Infection, SARS-CoV-2[MeSH Terms])) OR (Infection, SARS-CoV-2[Title/Abstract])) OR (SARS-CoV-2 Infections[MeSH Terms])) OR (SARS-CoV-2 Infections[Title/Abstract])) OR (2019 Novel Coronavirus Disease[MeSH Terms])) OR (2019 Novel Coronavirus Disease[Title/Abstract])) OR (2019 Novel Coronavirus Infection[MeSH Terms])) OR (2019 Novel Coronavirus Infection[Title/Abstract])) OR (2019-nCoV Disease[MeSH Terms])) OR (2019-nCoV Disease[Title/Abstract])) OR (2019-nCoV Diseases[MeSH Terms])) OR (2019-nCoV Diseases[Title/Abstract])) OR (Disease, 2019-nCoV[MeSH Terms])) OR (Disease, 2019-nCoV[Title/Abstract])) OR (COVID-19 Virus Infection[MeSH Terms])) OR (COVID-19 Virus Infection[Title/Abstract])) OR (COVID-19 Virus Infections[MeSH Terms])) OR (COVID-19 Virus Infections[Title/Abstract])) OR (Infection, COVID-19 Virus[MeSH Terms])) OR (Infection, COVID-19 Virus[Title/Abstract])) OR (Virus Infection, COVID-19[MeSH Terms])) OR (Virus Infection, COVID-19[Title/Abstract])) OR (Coronavirus Disease 2019[MeSH Terms])) OR (Coronavirus Disease 2019[Title/Abstract])) OR (Disease 2019, Coronavirus[MeSH Terms])) OR (Disease 2019, Coronavirus[Title/Abstract])) OR (Coronavirus Disease-19[MeSH Terms])) OR (Coronavirus Disease-19[Title/Abstract])) OR (Severe Acute Respiratory Syndrome Coronavirus 2 Infection[MeSH Terms])) OR (Severe Acute Respiratory Syndrome Coronavirus 2 Infection[Title/Abstract])) OR (SARS Coronavirus 2 Infection[MeSH Terms])) OR (SARS Coronavirus 2 Infection[Title/Abstract])) OR (COVID-19 Virus Disease[MeSH Terms])) OR (COVID-19 Virus Disease[Title/Abstract])) OR (COVID-19 Virus Diseases[MeSH Terms])) OR (COVID-19 Virus Diseases[Title/Abstract])) OR (Disease, COVID-19 Virus[MeSH Terms])) OR (Disease, COVID-19 Virus[Title/Abstract])) OR (Virus Disease, COVID-19[MeSH Terms])) OR (Virus Disease, COVID-19[Title/Abstract])) OR (2019-nCoV Infection[MeSH Terms])) OR (2019-nCoV Infection[Title/Abstract])) OR (2019-nCoV Infections[MeSH Terms])) OR (2019-nCoV Infections[Title/Abstract])) OR (Infection, 2019-nCoV[MeSH Terms])) OR (Infection, 2019-nCoV[Title/Abstract])) OR (COVID-19 Pandemic[MeSH Terms])) OR (COVID-19 Pandemic[Title/Abstract])) OR (Pandemic, COVID-19[MeSH Terms])) OR (Pandemic, COVID-19[Title/Abstract])) OR (COVID-19 Pandemics[MeSH Terms])) OR (COVID-19 Pandemics[Title/Abstract])) OR (SARS Coronavirus 2[MeSH Terms])) OR (SARS Coronavirus 2[Title/Abstract])) OR (Coronavirus 2, SARS[MeSH Terms])) OR (Coronavirus 2, SARS[Title/Abstract])) OR (Coronavirus Disease 2019 Virus[MeSH Terms])) OR (Coronavirus Disease 2019 Virus[Title/Abstract])) OR (2019 Novel Coronavirus[MeSH Terms])) OR (2019 Novel Coronavirus[Title/Abstract])) OR (2019 Novel Coronaviruses[MeSH Terms])) OR (2019 Novel Coronaviruses[Title/Abstract])) OR (Coronavirus, 2019 Novel[MeSH Terms])) OR (Coronavirus, 2019 Novel[Title/Abstract])) OR (Novel Coronavirus, 2019[MeSH Terms])) OR (Novel Coronavirus, 2019[Title/Abstract])) OR (SARS-CoV-2 Virus[MeSH Terms])) OR (SARS-CoV-2 Virus[Title/Abstract])) OR (SARS-CoV-2 Viruses[MeSH Terms])) OR (SARS-CoV-2 Viruses[Title/Abstract])) OR (Virus, SARS-CoV-2[MeSH Terms])) OR (Virus, SARS-CoV-2[Title/Abstract])) OR (2019-nCoV[MeSH Terms])) OR (2019-nCoV[Title/Abstract])) OR (COVID-19 Virus[MeSH Terms])) OR (COVID-19 Virus[Title/Abstract])) OR (COVID-19 Viruses[MeSH Terms])) OR (COVID-19 Viruses[Title/Abstract])) OR (Virus, COVID-19[MeSH Terms])) OR (Virus, COVID-19[Title/Abstract])) OR (Wuhan Coronavirus[MeSH Terms])) OR (Wuhan Coronavirus[Title/Abstract])) OR (Coronavirus, Wuhan[MeSH Terms])) OR (Coronavirus, Wuhan[Title/Abstract])) OR (Severe Acute Respiratory Syndrome Coronavirus 2[MeSH Terms])) OR (Severe Acute Respiratory Syndrome Coronavirus 2[Title/Abstract])) AND ((((((((((((((((((((((((((((((((((((((((Diabetes Mellitus[MeSH Terms]) OR (Diabetes Mellitus[Title/Abstract])) OR (Diabetes[Title/Abstract])) OR (Diabetic[Title/Abstract])) OR (Diabetes Mellitus, Type 1[MeSH Terms])) OR (Diabetes Mellitus, Type 1[Title/Abstract])) OR (Diabetes Mellitus, Insulin-Dependent[Title/Abstract])) OR (Diabetes Mellitus, Insulin Dependent[Title/Abstract])) OR (Insulin-Dependent Diabetes Mellitus[Title/Abstract])) OR (Diabetes Mellitus, Juvenile-Onset[Title/Abstract])) OR (Diabetes Mellitus, Juvenile Onset[Title/Abstract])) OR (Juvenile-Onset Diabetes Mellitus[Title/Abstract])) OR (Type 1 Diabetes Mellitus[Title/Abstract])) OR (Diabetes Mellitus, Insulin-Dependent, 1[Title/Abstract])) OR (Insulin-Dependent Diabetes Mellitus 1[Title/Abstract])) OR (Type 1 Diabetes[Title/Abstract])) OR (Diabetes, Type 1[Title/Abstract])) OR (Diabetes Mellitus, Type I[Title/Abstract])) OR (Diabetes, Autoimmune[Title/Abstract])) OR (Autoimmune Diabetes[Title/Abstract])) OR (IDDM[Title/Abstract])) OR (T1DM[Title/Abstract])) OR (Diabetes Mellitus, Type 2[MeSH Terms])) OR (Diabetes Mellitus, Type 2[Title/Abstract])) OR (Diabetes Mellitus, Noninsulin-Dependent[Title/Abstract])) OR (Diabetes Mellitus, Non Insulin Dependent[Title/Abstract])) OR (Diabetes Mellitus, Non-Insulin-Dependent[Title/Abstract])) OR (Non-Insulin-Dependent Diabetes Mellitus[Title/Abstract])) OR (Diabetes Mellitus, Type II[Title/Abstract])) OR (NIDDM[Title/Abstract])) OR (Diabetes Mellitus, Noninsulin Dependent[Title/Abstract])) OR (Type 2 Diabetes Mellitus[Title/Abstract])) OR (Noninsulin-Dependent Diabetes Mellitus[Title/Abstract])) OR (Noninsulin Dependent Diabetes Mellitus[Title/Abstract])) OR (Type 2 Diabetes[Title/Abstract])) OR (Diabetes, Type 2[Title/Abstract])) OR (Diabetes Mellitus, Adult-Onset[Title/Abstract])) OR (Adult-Onset Diabetes Mellitus[Title/Abstract])) OR (Diabetes Mellitus, Adult Onset[Title/Abstract])) OR (T2DM[Title/Abstract]))) AND (((((((((((((((((((((((((((((((((((((((Hospital*[MeSH Terms]) OR (Hospital*[Title/Abstract])) OR (Patient Admission[MeSH Terms])) OR (Patient Admission[Title/Abstract])) OR (Admission, Patient[Title/Abstract])) OR (Admissions, Patient[Title/Abstract])) OR (Patient Admissions[Title/Abstract])) OR (Stay, Hospital[Title/Abstract])) OR (Stays, Hospital[Title/Abstract])) OR (Intensive Care Units[MeSH Terms])) OR (Intensive Care Units[Title/Abstract])) OR (Intensive Care Unit[Title/Abstract])) OR (Unit, Intensive Care[Title/Abstract])) OR (ICU Intensive Care Units[Title/Abstract])) OR (ICU admission[Title/Abstract])) OR (ICU[Title/Abstract])) OR (Critical Care[MeSH Terms])) OR (Critical Care[Title/Abstract])) OR (Care, Critical[Title/Abstract])) OR (Intensive Care[Title/Abstract])) OR (Care, Intensive[Title/Abstract])) OR (Critical Care Unit[Title/Abstract])) OR (Critical Care Units[Title/Abstract])) OR (Intensive Therapy Unit[Title/Abstract])) OR (Intensive Treatment Unit[Title/Abstract])) OR (Critical Care Outcomes[MeSH Terms])) OR (Critical Care Outcomes[Title/Abstract])) OR (Care Outcome, Critical[Title/Abstract])) OR (Care Outcomes, Critical[Title/Abstract])) OR (Critical Care Outcome[Title/Abstract])) OR (Outcome, Critical Care[Title/Abstract])) OR (Outcomes, Critical Care[Title/Abstract])) OR (Mortal*[MeSH Terms])) OR (Mortal*[Title/Abstract])) OR (Death[Title/Abstract])) OR (Death Rate[Title/Abstract])) OR (Death Rates[Title/Abstract])) OR (Mortality Rate[Title/Abstract])) OR (Rate, Mortality[Title/Abstract]))

**#6:** (diabetes AND hospitalization) AND LitCFORECASTING[filter]

**#7:** (diabetes AND intensive care units) AND LitCFORECASTING[filter]

**#8:** (diabetes AND critical care) AND LitCFORECASTING[filter]

**#9:** (diabetes AND critical care outcomes) AND LitCFORECASTING[filter]

**#10:** (diabetes AND mortality) AND LitCFORECASTING[filter]

**#11:** (diabetes AND patient admission) AND LitCFORECASTING[filter]

**#12:** (diabetes mellitus AND hospitalization) AND LitCFORECASTING[filter]

**#13:** (diabetes mellitus AND intensive care units) AND LitCFORECASTING[filter]

**#14:** (diabetes mellitus AND critical care) AND LitCFORECASTING[filter]

**#15:** (diabetes mellitus AND critical care outcomes) AND LitCFORECASTING[filter]

**#16:** (diabetes mellitus AND mortality) AND LitCFORECASTING[filter]

**#17:** (diabetes mellitus AND patient admission) AND LitCFORECASTING[filter]

**#18:** (diabetes mellitus type 1 AND hospitalization) AND LitCFORECASTING[filter]

**#19:** (diabetes mellitus type 1 AND intensive care units) AND LitCFORECASTING[filter]

**#20:** (diabetes mellitus type 1 AND critical care) AND LitCFORECASTING[filter]

**#21:** (diabetes mellitus type 1 AND mortality) AND LitCFORECASTING[filter]

**#22:** (diabetes mellitus type 1 AND patient admission) AND LitCFORECASTING[filter]

**#23:** (diabetes mellitus type 2 AND hospitalization) AND LitCFORECASTING[filter]

**#24:** (diabetes mellitus type 2 AND intensive care units) AND LitCFORECASTING[filter]

**#26:** (diabetes mellitus type 2 AND critical care) AND LitCFORECASTING[filter]

**#27:** (diabetes mellitus type 2 AND mortality) AND LitCFORECASTING[filter]

**#28:** (diabetes mellitus type 2 AND patient admission) AND LitCFORECASTING[filter]

**#29:** ((((((((((((((((((((((diabetes AND hospitalization) AND LitCFORECASTING[filter]) OR ((diabetes AND intensive care units) AND LitCFORECASTING[filter])) OR ((diabetes AND critical care) AND LitCFORECASTING[filter])) OR ((diabetes AND critical care outcomes) AND LitCFORECASTING[filter])) OR ((diabetes AND mortality) AND LitCFORECASTING[filter])) OR ((diabetes AND patient admission) AND LitCFORECASTING[filter])) OR ((diabetes mellitus AND hospitalization) AND LitCFORECASTING[filter])) OR ((diabetes mellitus AND intensive care units) AND LitCFORECASTING[filter])) OR ((diabetes mellitus AND critical care) AND LitCFORECASTING[filter])) OR ((diabetes mellitus AND critical care outcomes) AND LitCFORECASTING[filter])) OR ((diabetes mellitus AND mortality) AND LitCFORECASTING[filter])) OR ((diabetes mellitus AND patient admission) AND LitCFORECASTING[filter])) OR ((diabetes mellitus type 1 AND hospitalization) AND LitCFORECASTING[filter])) OR ((diabetes mellitus type 1 AND intensive care units) AND LitCFORECASTING[filter])) OR ((diabetes mellitus type 1 AND critical care) AND LitCFORECASTING[filter])) OR ((diabetes mellitus type 1 AND mortality) AND LitCFORECASTING[filter])) OR ((diabetes mellitus type 1 AND patient admission) AND LitCFORECASTING[filter])) OR ((diabetes mellitus type 2 AND hospitalization) AND LitCFORECASTING[filter])) OR ((diabetes mellitus type 2 AND intensive care units) AND LitCFORECASTING[filter])) OR ((diabetes mellitus type 2 AND critical care) AND LitCFORECASTING[filter])) OR ((diabetes mellitus type 2 AND mortality) AND LitCFORECASTING[filter])) OR ((diabetes mellitus type 2 AND patient admission) AND LitCFORECASTING[filter])

**#30:** (((((((((((((((((((((((((((((((((((((((((((((((((((((((((((((((((((((((((((((((((((((((((((((COVID-19[MeSH Terms]) OR (COVID-19[Title/Abstract])) OR (SARS-CoV-2[MeSH Terms])) OR (SARS-CoV-2[Title/Abstract])) OR (SARS-CoV-2 Infection[MeSH Terms])) OR (SARS-CoV-2 Infection[Title/Abstract])) OR (Infection, SARS-CoV-2[MeSH Terms])) OR (Infection, SARS-CoV-2[Title/Abstract])) OR (SARS-CoV-2 Infections[MeSH Terms])) OR (SARS-CoV-2 Infections[Title/Abstract])) OR (2019 Novel Coronavirus Disease[MeSH Terms])) OR (2019 Novel Coronavirus Disease[Title/Abstract])) OR (2019 Novel Coronavirus Infection[MeSH Terms])) OR (2019 Novel Coronavirus Infection[Title/Abstract])) OR (2019-nCoV Disease[MeSH Terms])) OR (2019-nCoV Disease[Title/Abstract])) OR (2019-nCoV Diseases[MeSH Terms])) OR (2019-nCoV Diseases[Title/Abstract])) OR (Disease, 2019-nCoV[MeSH Terms])) OR (Disease, 2019-nCoV[Title/Abstract])) OR (COVID-19 Virus Infection[MeSH Terms])) OR (COVID-19 Virus Infection[Title/Abstract])) OR (COVID-19 Virus Infections[MeSH Terms])) OR (COVID-19 Virus Infections[Title/Abstract])) OR (Infection, COVID-19 Virus[MeSH Terms])) OR (Infection, COVID-19 Virus[Title/Abstract])) OR (Virus Infection, COVID-19[MeSH Terms])) OR (Virus Infection, COVID-19[Title/Abstract])) OR (Coronavirus Disease 2019[MeSH Terms])) OR (Coronavirus Disease 2019[Title/Abstract])) OR (Disease 2019, Coronavirus[MeSH Terms])) OR (Disease 2019, Coronavirus[Title/Abstract])) OR (Coronavirus Disease-19[MeSH Terms])) OR (Coronavirus Disease-19[Title/Abstract])) OR (Severe Acute Respiratory Syndrome Coronavirus 2 Infection[MeSH Terms])) OR (Severe Acute Respiratory Syndrome Coronavirus 2 Infection[Title/Abstract])) OR (SARS Coronavirus 2 Infection[MeSH Terms])) OR (SARS Coronavirus 2 Infection[Title/Abstract])) OR (COVID-19 Virus Disease[MeSH Terms])) OR (COVID-19 Virus Disease[Title/Abstract])) OR (COVID-19 Virus Diseases[MeSH Terms])) OR (COVID-19 Virus Diseases[Title/Abstract])) OR (Disease, COVID-19 Virus[MeSH Terms])) OR (Disease, COVID-19 Virus[Title/Abstract])) OR (Virus Disease, COVID-19[MeSH Terms])) OR (Virus Disease, COVID-19[Title/Abstract])) OR (2019-nCoV Infection[MeSH Terms])) OR (2019-nCoV Infection[Title/Abstract])) OR (2019-nCoV Infections[MeSH Terms])) OR (2019-nCoV Infections[Title/Abstract])) OR (Infection, 2019-nCoV[MeSH Terms])) OR (Infection, 2019-nCoV[Title/Abstract])) OR (COVID-19 Pandemic[MeSH Terms])) OR (COVID-19 Pandemic[Title/Abstract])) OR (Pandemic, COVID-19[MeSH Terms])) OR (Pandemic, COVID-19[Title/Abstract])) OR (COVID-19 Pandemics[MeSH Terms])) OR (COVID-19 Pandemics[Title/Abstract])) OR (SARS Coronavirus 2[MeSH Terms])) OR (SARS Coronavirus 2[Title/Abstract])) OR (Coronavirus 2, SARS[MeSH Terms])) OR (Coronavirus 2, SARS[Title/Abstract])) OR (Coronavirus Disease 2019 Virus[MeSH Terms])) OR (Coronavirus Disease 2019 Virus[Title/Abstract])) OR (2019 Novel Coronavirus[MeSH Terms])) OR (2019 Novel Coronavirus[Title/Abstract])) OR (2019 Novel Coronaviruses[MeSH Terms])) OR (2019 Novel Coronaviruses[Title/Abstract])) OR (Coronavirus, 2019 Novel[MeSH Terms])) OR (Coronavirus, 2019 Novel[Title/Abstract])) OR (Novel Coronavirus, 2019[MeSH Terms])) OR (Novel Coronavirus, 2019[Title/Abstract])) OR (SARS-CoV-2 Virus[MeSH Terms])) OR (SARS-CoV-2 Virus[Title/Abstract])) OR (SARS-CoV-2 Viruses[MeSH Terms])) OR (SARS-CoV-2 Viruses[Title/Abstract])) OR (Virus, SARS-CoV-2[MeSH Terms])) OR (Virus, SARS-CoV-2[Title/Abstract])) OR (2019-nCoV[MeSH Terms])) OR (2019-nCoV[Title/Abstract])) OR (COVID-19 Virus[MeSH Terms])) OR (COVID-19 Virus[Title/Abstract])) OR (COVID-19 Viruses[MeSH Terms])) OR (COVID-19 Viruses[Title/Abstract])) OR (Virus, COVID-19[MeSH Terms])) OR (Virus, COVID-19[Title/Abstract])) OR (Wuhan Coronavirus[MeSH Terms])) OR (Wuhan Coronavirus[Title/Abstract])) OR (Coronavirus, Wuhan[MeSH Terms])) OR (Coronavirus, Wuhan[Title/Abstract])) OR (Severe Acute Respiratory Syndrome Coronavirus 2[MeSH Terms])) OR (Severe Acute Respiratory Syndrome Coronavirus 2[Title/Abstract])) AND ((((((((((((((((((((((((((((((((((((((((Diabetes Mellitus[MeSH Terms]) OR (Diabetes Mellitus[Title/Abstract])) OR (Diabetes[Title/Abstract])) OR (Diabetic[Title/Abstract])) OR (Diabetes Mellitus, Type 1[MeSH Terms])) OR (Diabetes Mellitus, Type 1[Title/Abstract])) OR (Diabetes Mellitus, Insulin-Dependent[Title/Abstract])) OR (Diabetes Mellitus, Insulin Dependent[Title/Abstract])) OR (Insulin-Dependent Diabetes Mellitus[Title/Abstract])) OR (Diabetes Mellitus, Juvenile-Onset[Title/Abstract])) OR (Diabetes Mellitus, Juvenile Onset[Title/Abstract])) OR (Juvenile-Onset Diabetes Mellitus[Title/Abstract])) OR (Type 1 Diabetes Mellitus[Title/Abstract])) OR (Diabetes Mellitus, Insulin-Dependent, 1[Title/Abstract])) OR (Insulin-Dependent Diabetes Mellitus 1[Title/Abstract])) OR (Type 1 Diabetes[Title/Abstract])) OR (Diabetes, Type 1[Title/Abstract])) OR (Diabetes Mellitus, Type I[Title/Abstract])) OR (Diabetes, Autoimmune[Title/Abstract])) OR (Autoimmune Diabetes[Title/Abstract])) OR (IDDM[Title/Abstract])) OR (T1DM[Title/Abstract])) OR (Diabetes Mellitus, Type 2[MeSH Terms])) OR (Diabetes Mellitus, Type 2[Title/Abstract])) OR (Diabetes Mellitus, Noninsulin-Dependent[Title/Abstract])) OR (Diabetes Mellitus, Non Insulin Dependent[Title/Abstract])) OR (Diabetes Mellitus, Non-Insulin-Dependent[Title/Abstract])) OR (Non-Insulin-Dependent Diabetes Mellitus[Title/Abstract])) OR (Diabetes Mellitus, Type II[Title/Abstract])) OR (NIDDM[Title/Abstract])) OR (Diabetes Mellitus, Noninsulin Dependent[Title/Abstract])) OR (Type 2 Diabetes Mellitus[Title/Abstract])) OR (Noninsulin-Dependent Diabetes Mellitus[Title/Abstract])) OR (Noninsulin Dependent Diabetes Mellitus[Title/Abstract])) OR (Type 2 Diabetes[Title/Abstract])) OR (Diabetes, Type 2[Title/Abstract])) OR (Diabetes Mellitus, Adult-Onset[Title/Abstract])) OR (Adult-Onset Diabetes Mellitus[Title/Abstract])) OR (Diabetes Mellitus, Adult Onset[Title/Abstract])) OR (T2DM[Title/Abstract]))) AND (((((((((((((((((((((((((((((((((((((((Hospital*[MeSH Terms]) OR (Hospital*[Title/Abstract])) OR (Patient Admission[MeSH Terms])) OR (Patient Admission[Title/Abstract])) OR (Admission, Patient[Title/Abstract])) OR (Admissions, Patient[Title/Abstract])) OR (Patient Admissions[Title/Abstract])) OR (Stay, Hospital[Title/Abstract])) OR (Stays, Hospital[Title/Abstract])) OR (Intensive Care Units[MeSH Terms])) OR (Intensive Care Units[Title/Abstract])) OR (Intensive Care Unit[Title/Abstract])) OR (Unit, Intensive Care[Title/Abstract])) OR (ICU Intensive Care Units[Title/Abstract])) OR (ICU admission[Title/Abstract])) OR (ICU[Title/Abstract])) OR (Critical Care[MeSH Terms])) OR (Critical Care[Title/Abstract])) OR (Care, Critical[Title/Abstract])) OR (Intensive Care[Title/Abstract])) OR (Care, Intensive[Title/Abstract])) OR (Critical Care Unit[Title/Abstract])) OR (Critical Care Units[Title/Abstract])) OR (Intensive Therapy Unit[Title/Abstract])) OR (Intensive Treatment Unit[Title/Abstract])) OR (Critical Care Outcomes[MeSH Terms])) OR (Critical Care Outcomes[Title/Abstract])) OR (Care Outcome, Critical[Title/Abstract])) OR (Care Outcomes, Critical[Title/Abstract])) OR (Critical Care Outcome[Title/Abstract])) OR (Outcome, Critical Care[Title/Abstract])) OR (Outcomes, Critical Care[Title/Abstract])) OR (Mortal*[MeSH Terms])) OR (Mortal*[Title/Abstract])) OR (Death[Title/Abstract])) OR (Death Rate[Title/Abstract])) OR (Death Rates[Title/Abstract])) OR (Mortality Rate[Title/Abstract])) OR (Rate, Mortality[Title/Abstract]))) OR (((((((((((((((((((((((diabetes AND hospitalization) AND LitCFORECASTING[filter]) OR ((diabetes AND intensive care units) AND LitCFORECASTING[filter])) OR ((diabetes AND critical care) AND LitCFORECASTING[filter])) OR ((diabetes AND critical care outcomes) AND LitCFORECASTING[filter])) OR ((diabetes AND mortality) AND LitCFORECASTING[filter])) OR ((diabetes AND patient admission) AND LitCFORECASTING[filter])) OR ((diabetes mellitus AND hospitalization) AND LitCFORECASTING[filter])) OR ((diabetes mellitus AND intensive care units) AND LitCFORECASTING[filter])) OR ((diabetes mellitus AND critical care) AND LitCFORECASTING[filter])) OR ((diabetes mellitus AND critical care outcomes) AND LitCFORECASTING[filter])) OR ((diabetes mellitus AND mortality) AND LitCFORECASTING[filter])) OR ((diabetes mellitus AND patient admission) AND LitCFORECASTING[filter])) OR ((diabetes mellitus type 1 AND hospitalization) AND LitCFORECASTING[filter])) OR ((diabetes mellitus type 1 AND intensive care units) AND LitCFORECASTING[filter])) OR ((diabetes mellitus type 1 AND critical care) AND LitCFORECASTING[filter])) OR ((diabetes mellitus type 1 AND mortality) AND LitCFORECASTING[filter])) OR ((diabetes mellitus type 1 AND patient admission) AND LitCFORECASTING[filter])) OR ((diabetes mellitus type 2 AND hospitalization) AND LitCFORECASTING[filter])) OR ((diabetes mellitus type 2 AND intensive care units) AND LitCFORECASTING[filter])) OR ((diabetes mellitus type 2 AND critical care) AND LitCFORECASTING[filter])) OR ((diabetes mellitus type 2 AND mortality) AND LitCFORECASTING[filter])) OR ((diabetes mellitus type 2 AND patient admission) AND LitCFORECASTING[filter]))

**EMBASE:**

('coronavirus disease 2019'/exp OR '2019 novel coronavirus disease' OR '2019 novel coronavirus epidemic' OR '2019 novel coronavirus infection' OR '2019-ncov disease' OR '2019-ncov infection' OR 'covid' OR 'covid 19' OR 'covid 19 induced pneumonia' OR 'covid 2019' OR 'covid-10' OR 'covid-19' OR 'covid-19 induced pneumonia' OR 'covid-19 pneumonia' OR 'covid19' OR 'sars coronavirus 2 infection' OR 'sars coronavirus 2 pneumonia' OR 'sars-cov-2 disease' OR 'sars-cov-2 infection' OR 'sars-cov-2 pneumonia' OR 'sars-cov2 disease' OR 'sars-cov2 infection' OR 'sarscov2 disease' OR 'sarscov2 infection' OR 'wuhan coronavirus disease' OR 'wuhan coronavirus infection' OR 'coronavirus disease 2' OR 'coronavirus disease 2010' OR 'coronavirus disease 2019' OR 'coronavirus disease 2019 pneumonia' OR 'coronavirus disease-19' OR 'coronavirus infection 2019' OR 'ncov 2019 disease' OR 'ncov 2019 infection' OR 'new coronavirus pneumonia' OR 'novel coronavirus 2019 disease' OR 'novel coronavirus 2019 infection' OR 'novel coronavirus disease 2019' OR 'novel coronavirus infected pneumonia' OR 'novel coronavirus infection 2019' OR 'novel coronavirus pneumonia' OR 'paucisymptomatic coronavirus disease 2019' OR 'severe acute respiratory syndrome 2' OR 'severe acute respiratory syndrome 2 pneumonia' OR 'severe acute respiratory syndrome cov-2 infection' OR 'severe acute respiratory syndrome coronavirus 2 infection' OR 'severe acute respiratory syndrome coronavirus 2019 infection') AND ('diabetes mellitus'/de OR 'diabetes' OR 'diabetes mellitus' OR 'diabetic' OR 'non insulin dependent diabetes mellitus'/de OR 'niddm (non insulin dependent diabetes mellitus)' OR 't2dm' OR 'adult onset diabetes' OR 'adult onset diabetes mellitus' OR 'diabetes mellitus type 2' OR 'diabetes mellitus type ii' OR 'diabetes mellitus, maturity onset' OR 'diabetes mellitus, non insulin dependent' OR 'diabetes mellitus, non-insulin-dependent' OR 'diabetes mellitus, type 2' OR 'diabetes mellitus, type ii' OR 'diabetes type 2' OR 'diabetes type ii' OR 'diabetes, adult onset' OR 'dm 2' OR 'insulin independent diabetes' OR 'insulin independent diabetes mellitus' OR 'maturity onset diabetes' OR 'maturity onset diabetes mellitus' OR 'maturity onset diabetes of the young' OR 'niddm' OR 'non insulin dependent diabetes' OR 'non insulin dependent diabetes mellitus' OR 'non-insulin-dependent diabetes mellitus' OR 'noninsulin dependent diabetes' OR 'noninsulin dependent diabetes mellitus' OR 'type 2 diabetes' OR 'type 2 diabetes mellitus' OR 'type ii diabetes' OR 'type ii diabetes mellitus' OR 'insulin dependent diabetes mellitus'/de OR 't1dm' OR 'diabetes mellitus type 1' OR 'diabetes mellitus type i' OR 'diabetes mellitus, insulin dependent' OR 'diabetes mellitus, insulin-dependent' OR 'diabetes mellitus, juvenile onset' OR 'diabetes mellitus, type 1' OR 'diabetes mellitus, type i' OR 'diabetes type 1' OR 'diabetes type i' OR 'diabetes, juvenile' OR 'dm 1' OR 'early onset diabetes mellitus' OR 'iddm' OR 'insulin dependent diabetes' OR 'insulin dependent diabetes mellitus' OR 'insulin-dependent diabetes mellitus' OR 'juvenile diabetes' OR 'juvenile diabetes mellitus' OR 'juvenile onset diabetes' OR 'juvenile onset diabetes mellitus' OR 'labile diabetes mellitus' OR 'type 1 diabetes' OR 'type 1 diabetes mellitus' OR 'type i diabetes' OR 'type i diabetes mellitus') AND ('hospitalization'/de OR 'hospital stay' OR 'hospitalization' OR 'short stay hospitalization' OR 'intensive care unit'/de OR 'icu`s' OR 'close attention unit' OR 'critical care unit' OR 'general icu' OR 'intensive care department' OR 'intensive care unit' OR 'intensive care units' OR 'intensive therapy unit' OR 'intensive treatment unit' OR 'respiratory care unit' OR 'respiratory care units' OR 'special care unit' OR 'unit, intensive care' OR 'hospital admission'/de OR 'admission, hospital' OR 'admitting department, hospital' OR 'hospital admission' OR 'hospital admittance' OR 'hospital admitting department' OR 'hospital admitting service' OR 'hospital admitting unit' OR 'patient admission' OR 'critical care outcome'/de OR 'critical care outcome' OR 'critical care outcomes' OR 'intensive care outcome' OR 'intensive care outcomes' OR 'mortality'/de OR 'mortality' OR 'mortality model') AND ('observational study'/exp OR 'non experimental studies' OR 'non experimental study' OR 'nonexperimental studies' OR 'nonexperimental study' OR 'observation studies' OR 'observation study' OR 'observational studies' OR 'observational studies as topic' OR 'observational study' OR 'observational study as topic')

**OVID MEDLINE:**

(("COVID-19" or "SARS-CoV-2" or "SARS-CoV-2 Infection" or "Severe Acute Respiratory Syndrome Coronavirus 2" or "2019 Novel Coronavirus Disease" or "2019-nCoV Disease" or "COVID-19 Virus Infection" or "Coronavirus Disease 2019" or "COVID-19 Pandemic" or "SARS Coronavirus 2" or "Coronavirus Disease 2019 Virus" or "Novel Coronavirus, 2019" or "SARS-CoV-2 Virus" or "Coronavirus, Wuhan") and ("Diabetes Mellitus" or "Diabetes Mellitus, Type 1" or "Diabetes Mellitus, Type 2" or "Diabetes" or "Diabetes Mellitus, Insulin-Dependent" or "Type 1 Diabetes Mellitus" or "Diabetes Mellitus, Noninsulin-Dependent" or "Type 2 Diabetes" or "Type 2 Diabetes Mellitus" or "Insulin-Dependent Diabetes Mellitus" or "Diabetes Mellitus, Juvenile-Onset" or "Diabetes Mellitus, Juvenile Onset" or "Diabetes, Autoimmune" or "Autoimmune Diabetes" or "IDDM" or "Non-Insulin-Dependent Diabetes Mellitus" or "Diabetes Mellitus, Type II" or "NIDDM" or "Diabetes Mellitus, Adult-Onset" or "T1DM" or "T2DM" or "Diabetes Mellitus, Insulin Dependent" or "Juvenile-Onset Diabetes Mellitus" or "Diabetes Mellitus, Insulin-Dependent, 1" or "Type 1 Diabetes" or "Diabetes, Type 1" or "Diabetes Mellitus, Type I" or "Diabetes Mellitus, Non Insulin Dependent" or "Diabetes Mellitus, Non-Insulin-Dependent" or "Diabetes Mellitus, Noninsulin Dependent" or "Noninsulin-Dependent Diabetes Mellitus" or "Noninsulin Dependent Diabetes Mellitus" or "Diabetes, Type 2" or "Adult-Onset Diabetes Mellitus" or "Diabetes Mellitus, Adult Onset") and ("Hospital*" or "Patient Admission" or "Intensive Care Units" or "Critical Care" or "Critical Care Outcomes" or "Mortal*" or "Admission, Patient" or "Stay, Hospital" or "Unit, Intensive Care" or "ICU admission" or "ICU" or "Intensive Care" or "Intensive Therapy Unit" or "Intensive Treatment Unit" or "Death" or "Mortality Rate" or "Death Rate")).ab,kf,sh,ti.

**LILACS:**

(("COVID-19" OR "SARS-CoV-2" OR "SARS-CoV-2 Infection" OR "Severe Acute Respiratory Syndrome Coronavirus 2" OR "2019 Novel Coronavirus Disease" OR "2019-nCoV Disease" OR "COVID-19 Virus Infection" OR "Coronavirus Disease 2019" OR "COVID-19 Pandemic" OR "SARS Coronavirus 2" OR "Coronavirus Disease 2019 Virus" OR "Novel Coronavirus, 2019" OR "SARS-CoV-2 Virus" OR "Coronavirus, Wuhan")) AND (("Diabetes Mellitus" OR "Diabetes Mellitus, Type 1" OR "Diabetes Mellitus, Type 2" OR "Diabetes" OR "Diabetes Mellitus, Insulin-Dependent" OR "Type 1 Diabetes Mellitus" OR "Diabetes Mellitus, Noninsulin-Dependent" OR "Type 2 Diabetes" OR "Type 2 Diabetes Mellitus" OR "Insulin-Dependent Diabetes Mellitus" OR "Diabetes Mellitus, Juvenile-Onset" OR "Diabetes Mellitus, Juvenile Onset" OR "Diabetes, Autoimmune" OR "Autoimmune Diabetes" OR "IDDM" OR "Non-Insulin-Dependent Diabetes Mellitus" OR "Diabetes Mellitus, Type II" OR "NIDDM" OR "Diabetes Mellitus, Adult-Onset" OR "T1DM" OR "T2DM" OR "Diabetes Mellitus, Insulin Dependent" OR "Juvenile-Onset Diabetes Mellitus" OR "Diabetes Mellitus, Insulin-Dependent, 1" OR "Type 1 Diabetes" or "Diabetes, Type 1" OR "Diabetes Mellitus, Type I" OR "Diabetes Mellitus, Non Insulin Dependent" OR "Diabetes Mellitus, Non-Insulin-Dependent" OR "Diabetes Mellitus, Noninsulin Dependent" OR "Noninsulin-Dependent Diabetes Mellitus" OR "Noninsulin Dependent Diabetes Mellitus" OR "Diabetes, Type 2" OR "Adult-Onset Diabetes Mellitus" OR "Diabetes Mellitus, Adult Onset")) AND (("Hospital*" OR "Patient Admission" OR "Intensive Care Units" OR "Critical Care" OR "Critical Care Outcomes" OR "Mortal*" OR "Admission, Patient" OR "Stay, Hospital" OR "Unit, Intensive Care" OR "ICU admission" OR "ICU" OR "Intensive Care" OR "Intensive Therapy Unit" OR "Intensive Treatment Unit" OR "Death" OR "Mortality Rate" OR "Death Rate"))

**COVID-19 Global literature on coronavirus disease (World Health Organization):**

(("Diabetes Mellitus" OR "Diabetes Mellitus, Type 1" OR "Diabetes Mellitus, Type 2" OR "Diabetes" OR "Diabetes Mellitus, Insulin-Dependent" OR "Type 1 Diabetes Mellitus" OR "Diabetes Mellitus, Noninsulin-Dependent" OR "Type 2 Diabetes" OR "Type 2 Diabetes Mellitus" OR "Insulin-Dependent Diabetes Mellitus" OR "Diabetes Mellitus, Juvenile-Onset" OR "Diabetes Mellitus, Juvenile Onset" OR "Diabetes, Autoimmune" OR "Autoimmune Diabetes" OR "IDDM" OR "Non-Insulin-Dependent Diabetes Mellitus" OR "Diabetes Mellitus, Type II" OR "NIDDM" OR "Diabetes Mellitus, Adult-Onset" OR "T1DM" OR "T2DM" OR "Diabetes Mellitus, Insulin Dependent" OR "Juvenile-Onset Diabetes Mellitus" OR "Diabetes Mellitus, Insulin-Dependent, 1" OR "Type 1 Diabetes" OR "Diabetes, Type 1" OR "Diabetes Mellitus, Type I" OR "Diabetes Mellitus, Non Insulin Dependent" OR "Diabetes Mellitus, Non-Insulin-Dependent" OR "Diabetes Mellitus, Noninsulin Dependent" OR "Noninsulin-Dependent Diabetes Mellitus" OR "Noninsulin Dependent Diabetes Mellitus" OR "Diabetes, Type 2" OR "Adult-Onset Diabetes Mellitus" OR "Diabetes Mellitus, Adult Onset")) AND (("Hospital*" OR "Patient Admission" OR "Intensive Care Units" OR "Critical Care" OR "Critical Care Outcomes" OR "Mortal*" OR "Admission, Patient" OR "Stay, Hospital" OR "Unit, Intensive Care" OR "ICU admission" OR "ICU" OR "Intensive Care" OR "Intensive Therapy Unit" OR "Intensive Treatment Unit" OR "Death" OR "Mortality Rate" OR "Death Rate")) AND type_of_study:("risk_factors_studies" OR "prognostic_studies" OR "observational_studies")
